# Anti-inflammatory SARS-CoV-2 T cell immunity in asymptomatic seronegative Kenyan adults

**DOI:** 10.1101/2023.02.17.23286075

**Authors:** Taraz Samandari, Joshua Ongalo, Kim McCarthy, Richard K. Biegon, Philister Madiega, Anne Mithika, Joseph Orinda, Grace M. Mboya, Patrick Mwaura, Omu Anzala, Clayton Onyango, Fredrick O. Oluoch, Eric Osoro, Charles-Antoine Dutertre, Nicole Tan, Shou Kit Hang, Smrithi Hariharaputran, David C Lye, Amy Herman-Roloff, Nina Le Bert, Antonio Bertoletti

## Abstract

Antibodies are used to estimate prevalence of past infection. However, T cell responses against SARS-CoV-2 may more accurately define prevalence because SARS-CoV-2-specific antibodies wane. In November-December 2021, we studied serological and cellular immune responses in residents of rural Kenya who had not experienced any respiratory symptom nor had contact with COVID-19 cases. Among participants we detected anti-spike antibodies in 41.0% and T cell responses against ≥2 SARS-CoV-2 proteins in 82.5%, which implies that serosurveys underestimate SARS-CoV-2 prevalence in settings where asymptomatic infections prevail. Distinct from cellular immunity in European and Asian COVID-19 convalescents, strong T cell immunogenicity was observed against viral accessory proteins in these asymptomatic Africans, as well as a higher IL-10/IFN-γ ratio cytokine profile, suggesting that environmental or genetic factors modulate pro-inflammatory responses.

**Funding:** U.S. Centers for Disease Control and Prevention, Division of Global Health Protection. Singapore Ministry of Health’s National Medical Research Council under its COVID-19 Research Fund (COVID19RF3-0060, COVID19RF-001 and COVID19RF-008) and the Singapore Ministry of Health’s National Medical Research Council MOH-000019 (MOH-StaR17Nov-0001).

## INTRODUCTION

SARS-CoV-2 infection in Africa has been characterized by a less severe disease profile than what has been observed elsewhere^1,2^. The incidence of severe COVID-19 has been particularly low in Kenya^3,4^. Even though an undercount of deaths from COVID-19 cannot be firmly excluded^3,4^, Kenya’s National Emergency Operations Centre reported that ∼90% of infections were asymptomatic during the COVID-19 pandemic. This might be mainly explained by the country’s youthful population (i.e., median of 19 years for Kenya versus 38 for the USA), but other factors such as cross-reactive immunity induced by other coronaviruses^5^ or commensal microorganisms^6^, trained immunity stimulated by live vaccines (i.e., Bacille Calmette-Guérin (BCG) and live oral polio vaccines)^7,8^, or a downregulation of the inflammatory response via helminth co-infection^9^ could play mitigating roles.

The prevalence of past infection is classically measured using serological assays and in Kenya, by January-March 2021, blood donor anti-spike antibody seroprevalence ranged from 38% in rural western counties to 62% in the capital city, Nairobi^10^. However, it has been repeatedly shown that among asymptomatic SARS-CoV-2-infected individuals antibody levels are frequently low or absent whilst T cell responses remain detectable^11–13^. Furthermore, kinetic studies showed that memory T cells^14^ persist longer than antibodies^15^ in blood, implying that seroprevalence as an indicator may underestimate the extent of asymptomatic SARS-CoV-2 exposure.

Here, we studied SARS-CoV-2-specific humoral and cellular immune responses in individuals from Kenya who never reported any symptom of respiratory infection and who were not knowingly in contact with COVID-19 patients. We enlisted the guidance of locally-resident community healthcare workers to identify study participants residing in rural areas of the counties Kisumu and Elgeyo Marakwet, two regions of Kenya that by December 7, 2021, had reported 569 and 94 cases of COVID-19 per 100,000 population, respectively. Samples collected during November-December 2021 were studied in parallel for the presence of spike-specific antibodies and for T cells specific for SARS-CoV-2 structural (membrane, nucleoprotein, spike) and accessory (ORF3a, ORF7, ORF8) proteins utilizing different methods of T cell characterization. Until now, SARS-CoV-2-specific T cells, which have been hypothesized to play a major role in the control of disease severity^16^, have only been examined in African populations with convalescent COVID-19^17,18^, while T cell response characteristics in asymptomatic Africans have never been studied.

### High frequency of multi-specific T cell responses in asymptomatic individuals

Study participants with no history of respiratory illness (cough, shortness of breath, fever, or sinus congestion) since December 2019, no contact with known cases of COVID-19 and no history of COVID-19 vaccination were recruited from Elgeyo Marakwet (n=40), and Kisumu counties (n=40; Figure 1A-B). Among participants, 42/80 (53%) were female and 65/80 (81%) were under the age of 50 years; 10/63 (16%) were HIV infected, the remainder (n=17) having declined HIV testing, and 5/80 (6%) were hypertensive (Table 1). All had negative nasal swab tests for SARS-CoV-2 by polymerase chain reaction. Antibodies specific for spike-antigens were first tested using a surrogate neutralizing antibody test (GenScript cPass)^19^ and anti-IgG-SARS-CoV-2 tests (InBios-Euroimmun two-step assay) measuring the level of IgG against the S1 region of the spike protein. Surrogate neutralizing antibodies were detected in 16/40 (40%) of participants from Elgeyo Marakwet and in 18/40 (43%) of participants from Kisumu (Figure 1C). Similar proportions were observed using the alternative InBios-Euroimmun assays (Figure 1C). To exclude that a primary infection with SARS-CoV-2 variants of concern (Delta, Omicron) might have induced spike specific antibodies that do not cross-react with the Wuhan spike, we tested the presence of antibodies specific for the receptor-binding domain (RBD) region of Delta and Omicron. There was an almost complete concordance between the detection of antibodies against Wuhan and Delta spike RBD (Figure 1D). Only two individuals presented antibodies specific for Delta-spike RBD in the absence of antibodies against Wuhan-spike RBD, while antibodies against Omicron-spike RBD were absent in all individuals except the two who presented the higher values of pseudo-neutralizing activity. This observation is consistent with evidence that the Delta SARS-CoV-2 variant was circulating in Kenya in the second quarter of 2021, while Omicron started to be detected in Kenya only in late November 2021^20^.

**Fig. 1:**
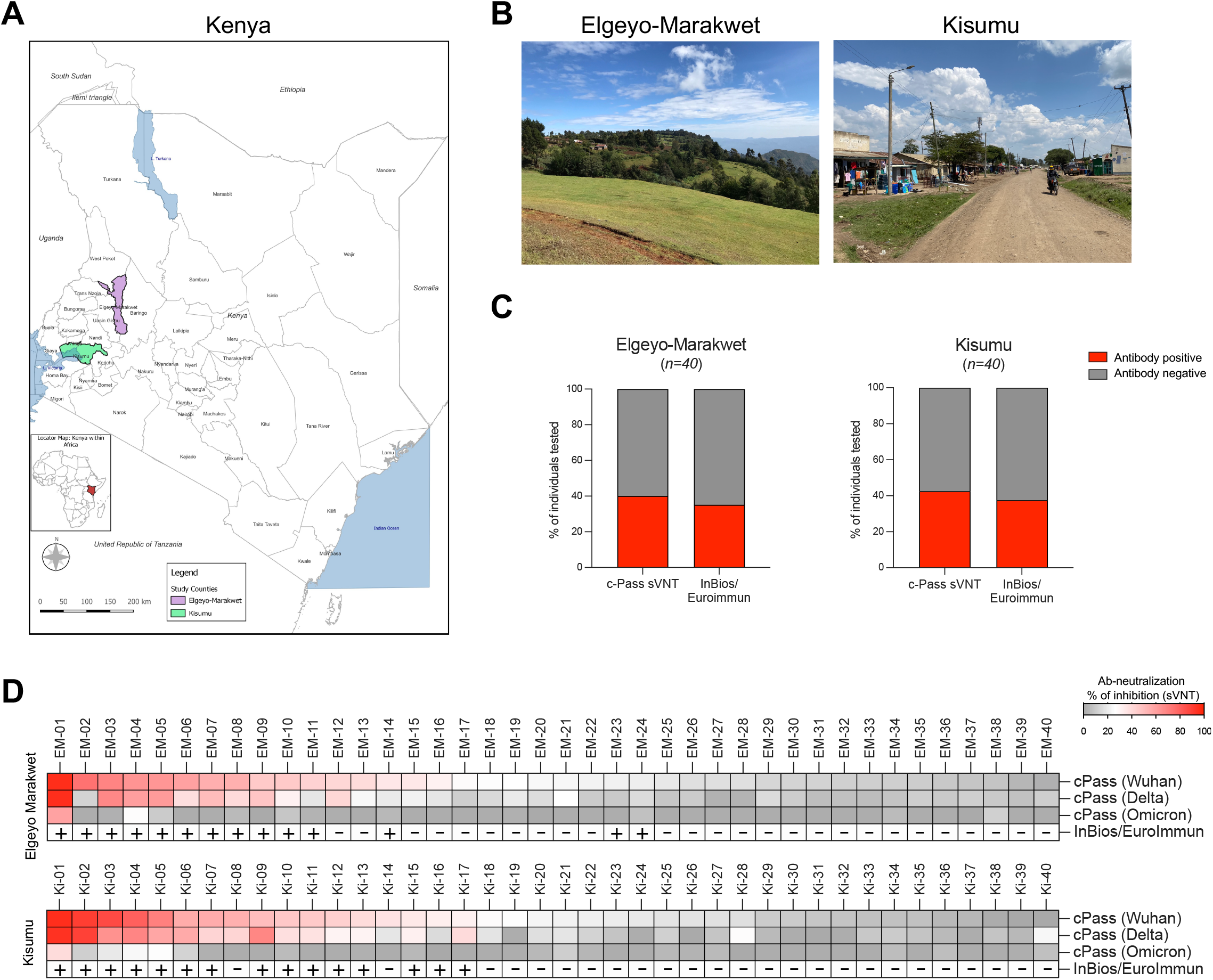
Frequency of SARS-CoV-2 seropositivity in asymptomatic individuals living in Elgeyo-Marakwet and Kisumu. (**A**) Individuals with no history of COVID-19 symptoms (including cough, shortness of breath, fever, or sinus congestion) and no contact with confirmed SARS-CoV-2 infected individuals were recruited in two counties of Kenya highlighted in purple and green: in (**B**) Elgeyo-Marakwet (n=40) and in Kisumu (n=40). (**C**) Sera of participants were analyzed with three antibody tests measuring Spike-specific antibodies: cPass measuring SARS-CoV-2 neutralizing antibodies with a surrogate virus neutralization test (sVNT); InBios SCoV-2 Detect IgG ELISA as an initial test followed by the Euroimmun anti-SARS-CoV-2 as a confirmatory test for participants who were positive by InBios. Percentage of participants positive for different tests are shown in red. (**D**) Antibody data are shown for the individual participants from Elgeyo Marakwet (above) and Kisumu (below). All participants were tested with the cPass sVNT against the spike protein of the first SARS-CoV-2 variant (Wuhan) and against the spike protein of SARS-CoV-2 variants Delta and Omicron. Test results form the InBios/EuroImmun tests are shown too.

**Table 1.** Demographic and clinical characteristics of asymptomatic Kenyan participants from Elgeyo Marakwet County and Kisumu County, 15 November-2 December, 2021

| **Characteristic** | **Kisumu** | **Elgeyo Marakwet** |
| --- | --- | --- |
| Age years, average (range) | 38 (18-73) | 37 (21-71) |
| Age ≥ 50 years, % (n/N) | 20% (8/40) | 18% (7/40) |
| Female, % (n/N) | 53% (21/40) | 53% (21/40) |
| Married, % (n/N) | 58% (23/40) | 70% (28/40) |
| Secondary educ, tech or university, % (n/N) | 38% (15/40) | 53% (21/40) |
| HIV-infected*, % (n/N) | 19% (7/36) | 11% (3/27) |
| Hypertension, % (n/N) | 3% (1/40) | 10% (4/40) |
| Diabetes, % (n/N) | 0% (0/40) | 0% (0/40) |

\* Some participants did not wish to be tested for HIV and did not share their HIV status. Of those who were HIV-infected the median CD4<sup>+</sup> cell count was 596 cells/ μL (range 363-1376).

T cell reactivity against SARS-CoV-2 structural (spike, nucleoprotein, membrane) and accessory (ORF3a, ORF7a/b, ORF8) proteins was tested by stimulating whole blood within 8 hours from sample collection using five distinct peptide pools (Fig 2A, left). After overnight incubation, IFN-γ and IL-2 were measured in the supernatants of the stimulated and unstimulated blood (Fig 2A, right) as previously reported^21^. We detected multi-specific anti-SARS-CoV-2 T cell responses (≥2 peptide pools) in 70% (28/40) of the Elgeyo Marakwet and in 95% (38/40) of the Kisumu groups (Fig 2B-C). Almost all individuals with positive anti-spike serology also showed cytokine responses against multiple SARS-CoV-2 proteins: 94% (15/16) of Elgeyo Marakwet and 100% (17/17) of the Kisumu groups. Furthermore, peptide-induced IFN-γ and IL-2 secretion were not only detectable in antibody seropositive individuals but also in the majority of seronegative ones: Elgeyo Marakwet 54% (13/24), Kisumu 91% (21/23, Figure 2B-D). Thus, only 27.5% of asymptomatic individuals from Elgeyo Marakwet and 5% from Kisumu were negative for both SARS-CoV-2 serology and T cell cytokine analysis (Fig 2D).

**Fig. 2.**
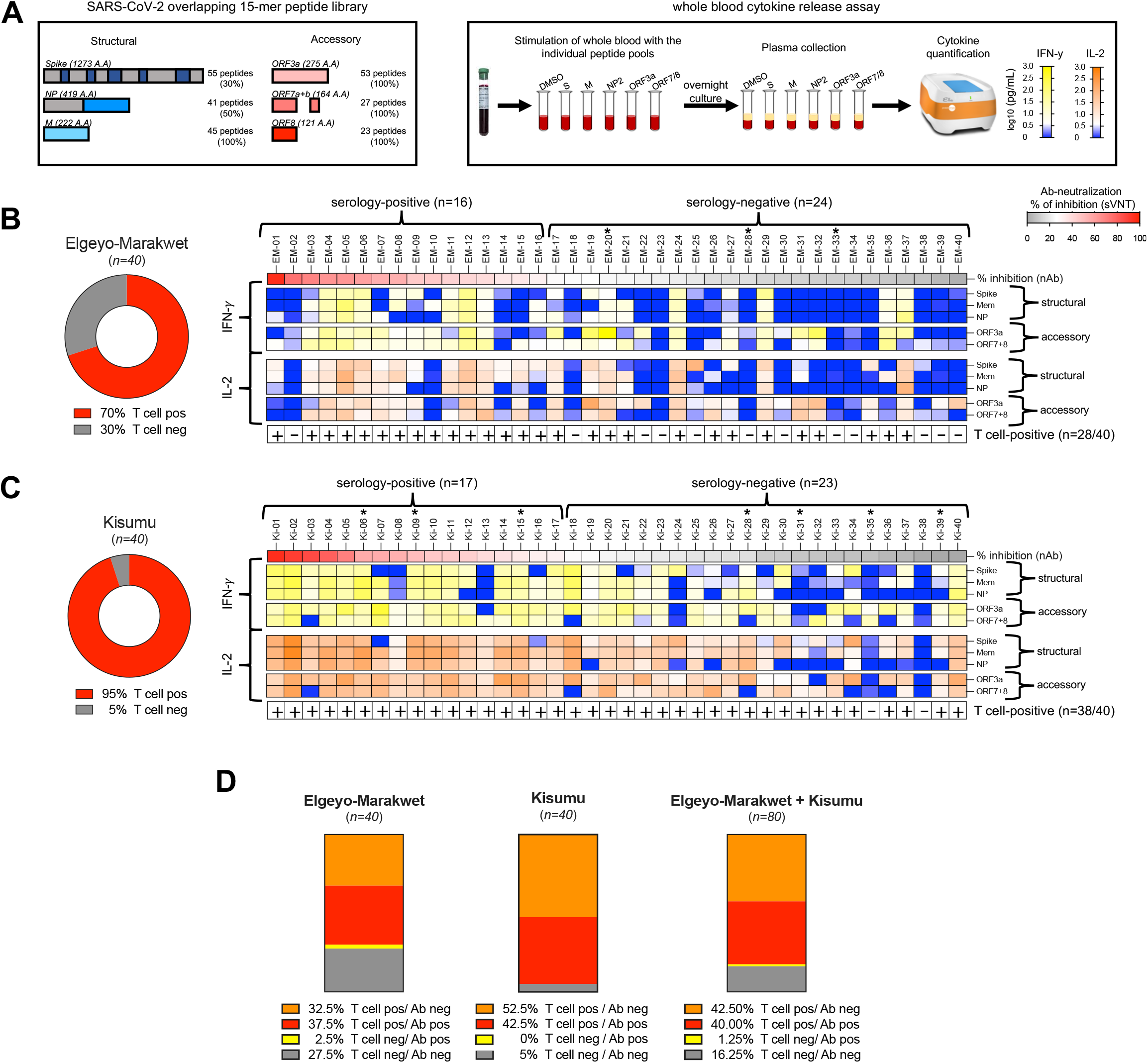
Frequency of T cells specific for different SARS-CoV-2 proteins in asymptomatic participants from Elgeyo-Marakwet and Kisumu. (**A**) Schematic representation of the five SARS-CoV-2-specific peptide pools containing 15-mer overlapping peptides spanning 30% of spike, entire sembrane (Mem) and 50% nucleoprotein (NP) and entire accessory proteins ORF3a and ORF7+8 that were used in 18h whole blood cultures. IFN- *μ* and IL-2 secreted in response to peptide stimulation were quantified in the plasma of the blood cultures. If peptide stimulation induced >5pg/ml of cytokines (IFN-*γ* and/or IL-2) above corresponding DMSO controls with two distinct peptide pools the individual was considered positive for SARS-CoV-2-specific T cells. Frequency of individuals from (**B**) Elgeyo-Marakwet and (**C**) Kisumu positive for SARS-CoV-2-specific T cells are shown in red (left). The heatmaps (right) show the levels of cytokines (yellow to blue: IFN-*γ* ; orange to blue: IL-2) released upon stimulation with the distinct peptide pools in each individual; HIV-positive individuals are indicated by *; samples considered positive for T cells are indicated by +. Participants are organized by level of neutralizing antibodies. (**D**) Percentage of individuals tested positive and/or negative for SARS-CoV-2-specific T cells and neutralizing antibodies.

Of note, some HIV-infected individuals displayed robust production of IFN-γ and IL-2. The HIV status of participants is indicated with an asterisk in figures 2B and C; the median CD4+ cell count was 596 cells/μL, and the range was 363-1376 cells/μL.

### Pre- and post-pandemic detection of T cells specific for structural and accessory SARS-CoV-2 proteins

The observation that serologically negative asymptomatic individuals produced T cell cytokines after whole blood stimulation with peptides covering different SARS-CoV-2 proteins suggests that they possess virus-specific T cells primed by SARS-CoV-2 infection. Indeed, the simultaneous presence of T cells specific for multiple SARS-CoV-2 proteins is characteristic of previous infection^22^. However, since pre-pandemic cross-reactive T cells, usually specific for a single protein, have been observed in 40-70% of individuals worldwide ^6,23–25^, we performed ELISpot assays by exposing thawed pre-pandemic PBMC from Nairobi as well as post-pandemic PBMC from Kisumu and Elgeyo Marakwet to peptide mixtures covering the entire lengths of three structural (spike, nucleocapsid, membrane) and three accessory proteins (ORF3a, ORF7, ORF8) of SARS-CoV-2 (Fig 3A). None of the PBMC collected before 2019 demonstrated multi-specific T cell responses to SARS-CoV-2 (Fig 3B). Single responses (>5 spots x 10^6^) to a peptide mixture covering spike, membrane, ORF3a or ORF7 were detected in 31% (4/13) of volunteers. In contrast, the ELISpot assays performed with post-pandemic PBMC (collected at the same time as the whole blood for T cell assays between 15 November-2 December 2021) confirmed the presence of T cell reactivity against multiple SARS-CoV-2 proteins in the great majority of the asymptomatic participants from Elgeyo Marakwet and Kisumu (Fig 3C-D). As the viability of the PBMC after freezing and thawing was suboptimal (low viability and/or failed positive controls), only 75% (60/80) of the samples were analyzable. ELISpot data confirmed an almost identical pattern to the results obtained using the whole blood rapid cytokine assays. T cells activated by at least 2 distinct peptide mixtures were detected in 73% and 74% of PBMC, respectively, collected in Kisumu and Elgeyo Marakwet. Of note, the results obtained with whole blood assay and ELISpot were identical in the Elgeyo Marakwet group (74% by ELISpot versus 70% by whole blood) in which the viability of PBMC was optimal (38 out of 40 samples) while discrepancies in the frequency of positive responses in the Kisumu group (73% by ELISpot versus 90% by whole blood) were associated with the poor viability of some samples implying that handling of samples can alter T cell immunological results^26^.

**Fig. 3.**
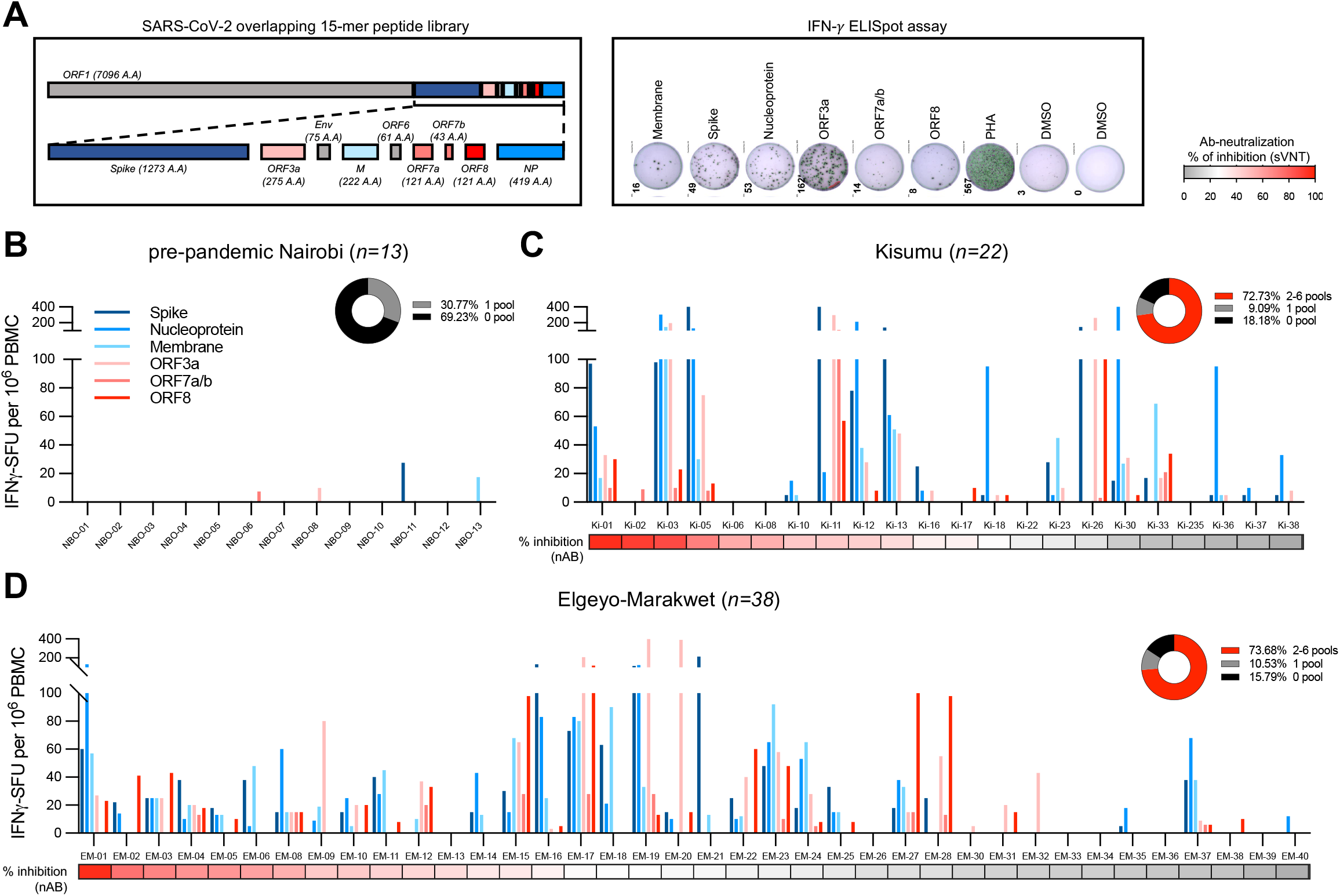
Frequency of T cells specific for different SARS-CoV-2 proteins analyzed by ELISpot in pre-pandemic samples from participants living in Nairobi, and from asymptomatic participants from Elgeyo-Marakwet and Kisumu collected in Dec 2021. (**A**) SARS-CoV-2 proteome organization; analyzed proteins are highlighted in color. PBMC were stimulated with 15-mer peptide pools overlapping by 10 amino acids covering the SARS-CoV-2 structural proteins spike, membrane and nucleoprotein and accessory proteins ORF3a, ORF7 and ORF8. IFN-*γ* secreting cells (SFU) in response to peptide stimulation were quantified by ELISpot assay. Frequency of IFN-*γ* secreting cells per 1 million PBMC is shown for each peptide pool in (**B**) pre-pandemic samples from Nairobi (n=13), and in samples collected from asymptomatic participants in December 2021 from (**C**) Kisumu (n=22) and (**D**) Elgeyo-Marakwet (n=38). Participants are organized by level of neutralizing antibodies (% inhibition by sVNT). (**B-D**) Inserted pie-charts show percentage of participants with responses to 2-6 peptide pools in red.

Finally, to unequivocally demonstrate that peptide mixtures were activating T cells, selected PBMC were stimulated with peptides, expanded in vitro, and then analyzed by flow cytometry for the presence of peptide-specific CD4+ or CD8+ T cells. Both SARS-CoV-2 peptide-specific CD4+ and CD8+ T cells were visualized, and their ability to recognize single peptides is shown (Fig S1).

### T cell immunodominance pattern to SARS-CoV-2 proteins

The utilization of peptide mixtures covering the whole length of the different structural (spike, membrane, nucleocapsid) and accessory (ORF3a, ORF7, ORF8) proteins in the ELISpot assays (Fig 4A) permitted the evaluation of the relative T cell immunogenicity of the proteins in 44 asymptomatic Kenyan participants. In individuals with multi-specific T cells, we calculated the percentage of T cells recognizing each protein. Bars in Figure 4B show the composition of the SARS-CoV-2 T cell response against different viral proteins for each individual. T cell immunogenicity was not proportional to the length of the protein tested. For example, despite spike consisting of 51% of the length of all proteins tested (Fig 4A), anti-spike specific T cells were the dominant T cell response in only 23% (10/44) of tested participants (Fig 4B). Interestingly, we noted a robust T cell response against the accessory proteins ORF3a and ORF8. Despite these proteins representing 15% of the length of all SARS-CoV-2 proteins tested (Fig 4A), ORF3a and ORF8 represented the dominant response in 30% (13/44) of asymptomatic individuals. The T cell immunodominance pattern in asymptomatic individuals living in rural Kenya was, however, distinctive to that in mild-moderately symptomatic COVID-19 convalescents (n=36) living in the urban environment of Singapore and tested 6 months after infection with identical methods and peptide pools (Fig 4C-E). Anti-spike activity clearly represented the dominant T cell response in Singaporean COVID-19 convalescents (Fig 4E). These results are consistent with observations by others who studied COVID-19 convalescents in the UK and USA^27–29^.

**Fig. 4.**
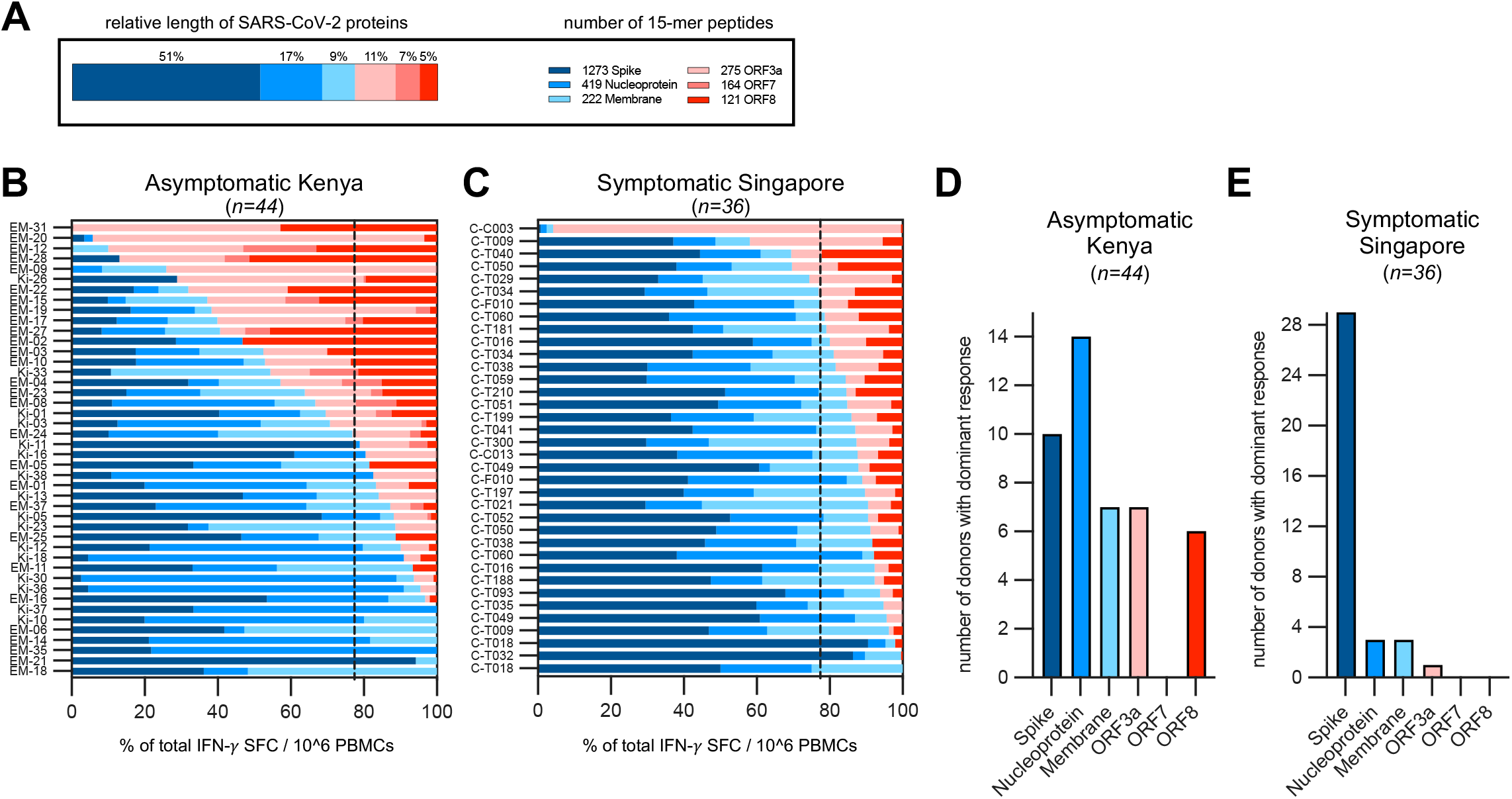
Immunodominance hierarchy of T cell responses to structural and accessory proteins of SARS-CoV-2. (**A**) Schematic representation of the relative length of the six different SARS-CoV-2 proteins tested and the number of 15-mer overlapping peptides covering the six proteins are indicated. (**B**) The composition of the SARS-CoV-2 T cell response in each responding asymptomatic participant from Kenya (n=44) is shown as a percentage of the total detected response (structural proteins = shades of blue; accessory proteins = shades of red). The dotted line represents the relative length of structural (77%) and accessory proteins (23%) tested. (**C**) The composition of the SARS-CoV-2 response in convalescent symptomatic COVID-19 patients from Singapore (n=36) is shown as a percentage of the total detected response. The number of participants with a dominant T cell response to the indicated SARS-CoV-2 proteins is shown for samples of asymptomatic participants from Kenya (**D**) and for symptomatic convalescent COVID-19 patients from Singapore (**E**).

### Functional dichotomy of T cells specific for structural and accessory proteins

IL-10 production by virus-specific T cells has been associated with reduced inflammation in respiratory viral infections^30,31^. Therefore, we compared the functional cellular immune response of asymptomatic participants from Kenya with COVID-19 convalescents from Singapore. We used unsupervised dimension reduction and clustering (UMAP) of the secretomes (IFN-γ, IL-2, and IL-10) of all peptide-stimulated samples (n=495) after subtraction of cytokine levels present in corresponding dimethyl sulfoxide controls. This showed that the secretion of cytokines classically produced by Th1 cells, IFN-γ and IL-2, was overlapping. In contrast, samples with high levels of the regulatory cytokine IL-10 formed a cluster with only partial intersection (Fig 5A). The overall secretomes from the three groups of participants differed (Fig 5B). The UMAP from Elgeyo Marakwet displays more samples with no or low level of cytokine release, consistent with Fig 2 showing that 30% of participants from this group had no SARS-CoV-2-specific cellular immunity. Many secretomes from both asymptomatic groups, Kisumu and Elgeyo Marakwet, cluster with high IL-10 levels, which is not seen for the secretomes from symptomatic COVID-19 convalescents from Singapore. Yet only samples from Kisumu and Singapore cluster on the UMAP with high levels of IFN-γ and IL-2.

**Fig. 5.**
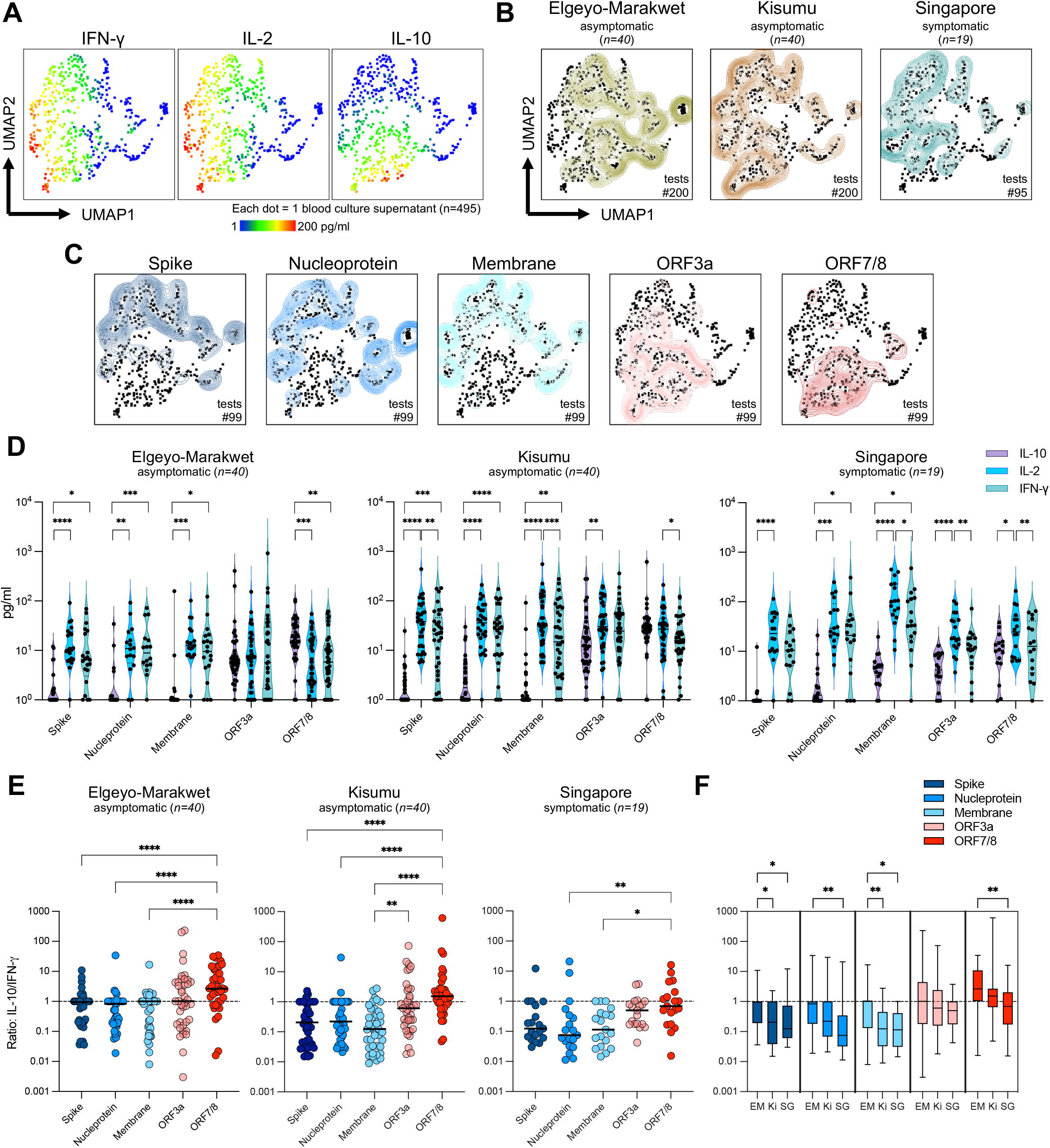
Cytokine secretion profile of whole blood from asymptomatic participants from Elgeyo-Marakwet and Kisumu and symptomatic convalescents from Singapore stimulated with SARS-CoV-2-peptide pools. Whole blood was stimulated with SARS-CoV-2-peptide pools overnight and the cytokine secretion profile (IFN-*γ*, IL-2 and IL-10) was analyzed using unsupervised clustering algorithm UMAP. (**A**) UMAP plots with cytokine secretion heatmaps. (**B**) Concatenated cytokine secretion profiles from asymptomatic participants from Elgeyo-Marakwet (left, green, 40 individuals, 200 tests), from Kisumu (middle, brown, 40 individuals, 200 tests) and from convalescent symptomatic COVID-19 patients from Singapore (right, blue, 19 individuals, 95 tests) overlaid on the global UMAP plot of all analyzed samples (black dots; each dot corresponds to one culture supernatant). (**C**) UMAP plots comparing the cytokine secretion profiles of whole blood from all individuals tested (n=99) stimulated with the 5 different SARS-CoV-2 peptide pools shown individually. (**D**) Violin plots showing the quantity of IL-10, IL-2 and IFN-*γ* detected in the different culture supernatants from asymptomatic participants from Elgeyo-Marakwet (left) and Kisumu (middle) and from symptomatic convalescents from Singapore (right). Friedmans test followed by Dunn’s multiple comparisons test; line = median. (**E**) Ratios of IL-10/IFN-*γ* quantities detected in the culture supernatants stimulated with the different peptide pools. (**F**) Ratios of IL-10/IFN-*γ* quantities detected in the culture supernatants stimulated with the different peptide pools are compared between the three cohorts; EM = Elgeyo-Marakwet; Ki = Kisumu; SG = Singapore). (**E-F**) Kruskal-Wallis test, followed by Dunn’s multiple comparisons test; line = median. P-values: <0.05 = *; <0.01 = **; <0.001 = ***; <0.0001 = ****.

Deconvolution of the secretion profiles in response to the individual peptide pools covering the different SARS-CoV-2 proteins showed distinctive profiles between structural and accessory proteins (Fig 5C). Cytokine profiles in response to accessory proteins clustered with high IL-10. A side-by-side comparison of the cytokines produced by each peptide pool in the three cohorts showed that while structural proteins produced a robust Th1 response (high IFN-γ/IL-2, low IL-10), accessory proteins induced similar quantities of IFN-γ and IL-10 (Fig 5D). In the participants from Elgeyo Marakwet, ORF7/8 triggered significantly higher secretion of the anti-inflammatory cytokine IL-10 than IFN-γ or IL-2.

The IL-10/IFN-γ ratio was highest for ORF7/8, followed by ORF3a in all three groups (Fig 5E). Comparison of the asymptomatic and convalescent groups revealed higher IL-10/IFN-γ ratios in samples from Elgeyo Marakwet, followed by Kisumu and lowest for Singapore in response to all the tested proteins. Differences in IL-10/IFN-γ ratios in samples from Elgeyo Marakwet and Kisumu were statistically significant only for T cell responses against spike and membrane. However, T cell responses against all the tested proteins except ORF3a showed statistically significantly higher IL-10/IFN-γ ratios in asymptomatic participants from Elgeyo Marakwet compared to Singaporean COVID-19 convalescents (Fig 5F).

## DISCUSSION

In a unique survey of both cellular and humoral immune responses against SARS-CoV-2 among asymptomatic Africans, we show that 78% of individuals living in two rural regions of Kenya who had no reported respiratory symptoms since December 2019 and who were never knowingly in contact with individuals infected with SARS-CoV-2, possessed broadly reactive T cells specific to multiple SARS-CoV-2 proteins. Sixty percent of these asymptomatic individuals lacked anti-spike antibodies – an antibody response that is more durable than the anti-nucleocapsid antibody^15^ – while among these seronegative participants 70% had multi-specific T cell responses. The simultaneous detection of a T cell response to different SARS-CoV-2 structural and accessory proteins in these individuals contrasts with the detection of T cell responses specific for single SARS-CoV-2 proteins from pre-pandemic samples.

In addition to strengthening the evidence that rates of asymptomatic SARS-CoV-2 infection were already very high in Kenya before the advent of the highly transmissible Omicron variant of concern, our results strongly suggest that measurement of virus-specific T cells constitutes a far more sensitive assay than antibodies to detect past coronavirus infections. This is consistent with the differential waning of antibody titers and T cell frequencies, particularly in individuals with no or minimal symptoms^15^ and with the detection of multi-specific SARS-CoV-2 T cell responses in the absence of antibodies in other studies of asymptomatic SARS-CoV-2 infection^11,12^. Furthermore, the persistence of virus-specific T cells over antibodies was also already observed in other coronavirus infections such as SARS-CoV-1^32^ and MERS^33,34^. Antibodies against SARS-CoV-1 are undetectable 2-3 years after infection^32^ while SARS-CoV-1-specific T cells are detectable up to at least 17 years after infection^23^. Similarly, T cell responses against MERS coronavirus were present in individuals with occupational exposure to camels in the absence of antibody responses^33,34^.

Despite such evidence, epidemiological assessments of the prevalence of SARS-CoV-2 have mainly utilized serologic assays since antibodies are easier to measure than T cells responses. However, methodologies to rapidly detect virus-specific T cell responses flourished during the COVID-19 pandemic^21,35–37^, including the use of whole blood which appeared to correlate with protection from SARS-CoV-2 infection in one study^37^. Here, we show directly that cytokine detection following whole blood stimulation with peptides can be implemented in locations within a few hours distance from a facility with biosafety level-1 cabinets. As these whole blood assays continue to be perfected for large scale use, they could eventually be applied routinely for public health surveillance of exposure to microbes that are known to elicit seronegative responses in asymptomatic individuals or in whom antibodies wane quickly.

If indeed there is a reduced antibody response to SARS-CoV-2 as observed in our cross-sectional study that ended in 2021, one may ask why there has been a steady increase in the seroprevalence of SARS-CoV-2 in Kenya and other countries of the African continent, reaching 90% or higher in many settings? Certainly, our study targeted asymptomatic individuals who are known to have reduced antibody positivity, whereas serosurveys encompass both symptomatic and asymptomatic persons. Nevertheless, there remains a wide discrepancy between the antibody and T cell responses. A clue comes from data in surveillance platforms in Kenya where a 5-10% polymerase chain reaction-positivity for SARS-CoV-2 has been observed among symptomatic individuals during inter-wave periods. We speculate that as SARS-CoV-2 transitioned from an epidemic to an endemic virus, continual re-infections have boosted the population’s antibody levels, especially as there has been very little promotion of non-pharmaceutical interventions in over a year. The fact that our participants were rural-based and had at least some degree of isolation from urban centers may have allowed time for their antibody levels to decline rather than get boosted through re-infection.

We observed that the Kisumu group of asymptomatic participants had a 95% multi-specific T cell response to SARS-CoV-2 proteins, while the proportion was lower (70%) for participants from Elgeyo Marakwet. We attribute this difference to the somewhat greater interaction of residents of peri-urban Kisumu with the city of Kisumu, which boasts a population of approximately 350,000, whereas the residents of Elgeyo Marakwet whom we enrolled, live a life distinctly more remote from urban centers.

These two rural communities have health and environmental characteristics that one may speculate influence the unique immune responses we observed. In both communities, the administration of a birth dose of BCG is routine, incident *Mycobacterium tuberculosis* disease and helminthic infections are commonplace (school children regularly receive anti-helminthics during biannual mass drug administration events, and persons living with HIV are routinely provided TB preventive therapy). While malaria is uncommon in Elgeyo Marakwet, it is endemic in Kisumu. Furthermore, while both groups often sleep in the same shelter as their livestock to prevent their theft, those in Elgeyo Marakwet typically own cows, prepare a fermented cow’s milk called *mursik* and exercise more due to the steep terrain at an altitude of approximately 1400 meters higher than Kisumu. Whether these environmental differences explain some of the unexpected features of SARS-CoV-2 T cell responses observed in asymptomatic rural Kenyans will require additional investigation.

We observed two distinctive immunologic characteristics among these asymptomatic rural Africans. The first was the observation of an unexpectedly strong T cell immunogenicity against the viral accessory proteins ORF3a and ORF8 that contrasts with SARS-CoV-2 T cells studied in convalescent urban Singaporeans and Western (UK and USA) patients who instead showed a clear dominance of spike-specific T cell responses^27–29^. Since the SARS-CoV-2 T cell repertoire can be modulated by exposure to commensal antigens^6^, the possibility that differing microbiomes can alter the SARS-CoV-2 T cell immunodominance cannot be excluded. An alternative hypothesis emerges from the kinetics of SARS-CoV-2 protein synthesis. Accessory molecules (ORF3a and ORF7a/b) are produced earlier after infection^38^, likely because they play a role in suppressing innate immunity in the infected cells^39^. Furthermore ORF3a, ORF7a/b^40^ and ORF8^41^ have been shown to reduce HLA-class I presentation. The hypothesis is that in the early phases of infection these proteins might be more immunogenic than structural proteins and in asymptomatic infections such abortive infections with limited virion production may predominate as compared with patients with symptomatic infections. Support for this hypothesis, is the finding of an early immune response against accessory proteins in acute COVID-19 patients^42^.

The second intriguing feature was the skewed IL-10 production of SARS-CoV-2 T cells detected preferentially in asymptomatic individuals living in rural Kenya but particularly in the rural communities of Elgeyo Marakwet. One interpretation is that specific environmental factors more common in rural areas of Kenya than in the city of Singapore altered the cytokine profile of SARS-CoV-2 T cells. The ability of helminths or mycobacteria (including BCG) to skew the cytokine production of immune cells has been well documented^43,44^ but other factors may also play a role. For example, in comparison to Kisumu participants, the relatively augmented levels of IL-10 production observed in Elgeyo Marakwet could be explained by the differences in lifestyle and diet or by the absence of malaria in this high altitude region. Exercise^45^ and fermented foods^46^ are associated with anti-inflammatory effects on the human immune response, while malaria has been shown to induce a Th1-like response^47^. It is interesting to point out that high levels of IL-10 were also detected in a study of asymptomatic infection occurring in Singapore at the beginning of the COVID-19 pandemic: in this case, the asymptomatic individuals were all young guest laborers who recently arrived from Bangladesh^12^. Whether the observed ability to produce more IL-10 by SARS-CoV-2 T cells can fully explain the asymptomatic profile of SARS-CoV-2 infection observed in these groups deserves further study.

Our study has several limitations. First, the study participants were not selected at random from local communities but were targeted by community healthcare workers due to the likelihood of their probable uninfected status and minimal contact with major urban centers. The small number of participants from these communities limits the generalizability of our findings to the rest of Kenya or perhaps even within these communities. Our comparison group for symptomatic convalescent individuals was hospitalized Singaporean patients with mild-moderate disease and not hospitalized individuals from these same communities. Finally, our pre-pandemic PBMC were from a healthy Nairobi cohort and not from either Kisumu or Elgeyo Marakwet because pre-pandemic PBMC from healthy volunteers in these counties were unavailable.

Our finding that cellular immune assays are more sensitive than antibody assays in detecting SARS-CoV-2 infection in an African population where asymptomatic infections have predominated, implies that seroprevalence surveys underestimate the spread of COVID-19. This observation, coupled with the knowledge that coronaviruses have the propensity to evolve into diseases with pandemic potential, should spur on the development of simple and scalable cellular immune assays to test populations for public health purposes. In order to see if our findings are generalizable, we encourage the public health research community to conduct similar studies on a much wider scale. We also observed that the T cell responses of asymptomatic residents of these rural Kenyan communities were unusually directed against SARS-CoV-2’s non-structural proteins and were skewed towards an anti-inflammatory response. These data support the call^1^ for a more in-depth analysis of the impact of environmental factors on the development of protective or pathological antiviral immunity against SARS-CoV-2 to understand COVID-19 pathogenesis not only in Africa but around the world.

## Data Availability

Data produced in the present study are available upon reasonable request to the authors

## Materials and Methods

### Experimental Design

We worked with local health departments and one facility each in Kisumu and Elgeyo Marakwet to identify a convenience sample of forty potential volunteers who lived in rural parts of each county. Participants enrolled in this cross-sectional study had to be at least 18 years of age and must not have received a COVID-19 vaccine. In addition, because the enrollees were intended to be individuals not exposed to SARS-CoV-2, they must have had no history of respiratory illness (cough, shortness of breath, fever, or sinus congestion) since December 31, 2019, no history of travel to or meeting with persons from high incidence counties such as Nairobi or Mombasa (i.e., having >500 cumulative cases per 100,000 as at January 2021) or history of incarceration since January 2020 and a negative SARS-CoV-2 PCR or antigen test at the time of enrolment. Kibigori and Tamu in Kisumu county are a 1.5 hour motorbike and bus ride away from Kisumu city; residents are engaged in subsistence agriculture and seasonal work on sugar plantations but could socialize with visiting city dwellers at local venues. Residents of Chesoi, Kapcherop, and Tambach in Elgeyo Marakwet are also engaged in subsistence farming but infrequently interact with dwellers of the nearest large city, Eldoret, a 2-hour bus ride away, located in a different county.

We enrolled participants in Kisumu between 15-18 November 2021 and in Elgeyo Marakwet between 29 November-2 December 2021. Up to 18.2 mL of blood was drawn from each volunteer, transported under controlled temperature, and a nasopharyngeal swab was collected to perform a PCR test for the presence of SARS-CoV-2. Additional consent was received to conduct an HIV rapid antibody test and – if accepted – to provide HIV prevention counselling.

Written informed consent was received from all participants in the cross-sectional survey. The Kenya Medical Research Institute’s (KEMRI) Scientific and Ethics Review Unit approved the protocol (KEMRI/RES/7/3/1, protocol #4186), the Kenya National Commission for Science, Technology and Innovation provided a permit (license no: NACOSTI/P/21/12171) and the United States’ Centers for Disease Control and Prevention relied upon the KEMRI approval (CDC #7353) as did the Washington State University Institutional Review Board.

As pre-pandemic frozen PBMC from either Kisumu or Elgeyo Marakwet were unavailable, we tested cross-reactivity against SARS-CoV-2 proteins in frozen PBMC collected from healthy volunteers in Nairobi between 19 October 2015 and 19 July 2016. These samples came from the SiVET study protocol, which received ethical approval from the Kenyatta National Hospital/University of Nairobi Ethics and Research Committee (P137/03/2015). Blood was also collected 6-months post SARS-CoV-2 infection from convalescent Singaporeans who had symptomatic COVID-19 in 2020 under COVID-19 PROTECT study group protocol which was approved by the National Healthcare Group (NHG) Domain Specific Review Board (DSRB) (number 2012/00917 and NHG DSRB E 2020/00091). All participants had provided written informed consent in accordance with the Declaration of Helsinki for Human Research.

### Laboratory procedures

#### Antibody assays

SARS-CoV-2 neutralizing antibodies were analyzed using a surrogate neutralization assay (cPass, GenScript Biotech, Singapore), which measures how neutralizing antibodies in the serum bind to HRP-labeld-SARS-CoV-2-Receptor Binding Domain (RBD) and prevent it from binding to the hACE2 protein (*19*). RBDs from three different SARS-CoV-2 variants were tested: ancestral Wuhan-strain, Delta and Omicron variants. A threshold of 30% inhibition was considered a positive result. Additionally, a two-step antibody detection test was conducted using the SCoV-2 Detect(tm) IgG ELISA (InBios International Inc., Seattle, Washington), as the initial test followed by the Euroimmun anti-SARS-CoV-2 ELISA (Euroimmun US, Mountain Lakes, NJ) as a confirmatory test for participants who were positive by InBios.

#### Cytokine release assays using whole blood

Details of this method are published elsewhere (*21*). 320 *μ*l of freshly drawn whole blood was stimulated with five distinct 15-mer peptide pools and controls (Fig 2A). After overnight culturing at 37°C with 5% CO2, the supernatant (plasma) was collected and frozen at -80°C for later shipment to our Singapore laboratory. Cytokine concentrations in the plasma were quantified using an Ella machine measuring IFN-γ, IL-2, and IL-10, following the manufacturer’s instructions (ProteinSimple). The level of cytokines present in the plasma of DMSO controls was subtracted from the corresponding peptide pool stimulated samples. Samples with cytokine quantities (IFN-γ, IL-2, IL-10) superior to 5pg/ml were considered positive. Since SARS-CoV-2 infection usually induces T cells specific not for single but multiple SARS-CoV-2 proteins (*22*), we scored as “T cell positive” only the individuals who exhibited a positive response to at least 2 peptide mixtures for each cytokine tested or a “multi-specific” response.

Subsequently, concentrations of each cytokine in all culture supernatants were transformed using the logicle transformation function, and UMAP was run using 15 nearest neighbors (*nn*), *min_dist* of 0.5, and Euclidean distance (*48*). The results obtained from UMAP analyses were incorporated as additional parameters and converted to FCS files, which were then loaded into FlowJo (BD Biosciences) to generate heatmaps of cytokine secretion on the reduced dimensions.

#### SARS-CoV-2 peptide-specific T cell quantification by ELISpot

The frequency of SARS-CoV-2 peptide-specific T cells was quantified as described previously (*42*). Briefly, cryopreserved PBMC that had been shipped to Singapore were thawed and stimulated with the following 15-mer peptide pools overlapping by 10 amino acids in ELISpot plates: structural (NP, Membrane, Spike) and accessory (ORF3a, ORF7, ORF8). The plates were then incubated with a human biotinylated IFN-γ detection antibody, followed by streptavidin–alkaline phosphatase (streptavidin-AP) and developed using the KPL BCIP/NBT phosphatase substrate (Seracare Life Sciences). The results were expressed as spot-forming cells (SFC) per 10^6^ PBMCs.

#### Cell culture for T cell expansion

T cell lines were generated as follows: 20% of PBMCs were pulsed with 10 µg/ml of the overlapping SARS-CoV-2 peptides for 1 h at 37°C, washed, and cocultured with the remaining cells in AIM-V medium (Gibco, Thermo Fisher Scientific) supplemented with 2% AB human serum (Gibco, Thermo Fisher Scientific). T cell lines were cultured for 10 d in the presence of 20 U/ml of recombinant IL-2 (R&D Systems).

#### Flow cytometry

PBMCs were stimulated with peptide pools and expanded in vitro for 10 days, as described before. Expanded T cell lines were stimulated for 5 h at 37°C with or without SARS-CoV-2 peptide pools (2 µg/ml). After 1 h, 10 µg/ml brefeldin A (Sigma-Aldrich) and 1× monensin (BioLegend) were added. Cells were stained with the yellow LIVE/DEAD fixable dead cell stain kit (Invitrogen) and surface marker: anti-CD3 (SK7 or OKT3; BioLegend), anti-CD4 (SK3), anti-CD8 (SK1). Cells were subsequently fixed and permeabilized using the Cytofix/Cytoperm kit (BD Biosciences) and stained with anti–IFN-γ (25723; R&D Systems) and anti–TNF-α (MAb11) antibodies and analyzed on a CytoFLEX (Beckman Coulter). Data were analyzed by FlowJo (BD Biosciences). Antibodies were purchased from BD Biosciences unless otherwise stated.

#### Statistical analysis

All tests are described in the figure legends.

## Supplementary Figures

fig. S1 Phenotypic analysis of T cells specific to single SARS-CoV-2 peptides

PBMC were in vitro expanded with peptide pool ORF3a for 10 days and responses to individual peptides were identified stimulating the expanded cells with a peptide matrix strategy using a ELISpot assay. Responses to single peptides were then confirmed by flow cytometry analysis. (A) Flow cytometry gating strategy. (B) Participant EM-06 with CD8 T cell responses specific to two different peptides within ORF3a. (C) Participants EM-32 and EM-06 with CD4 T cell responses specific to two peptides within ORF3a.

**Supplementary Fig. 1:**
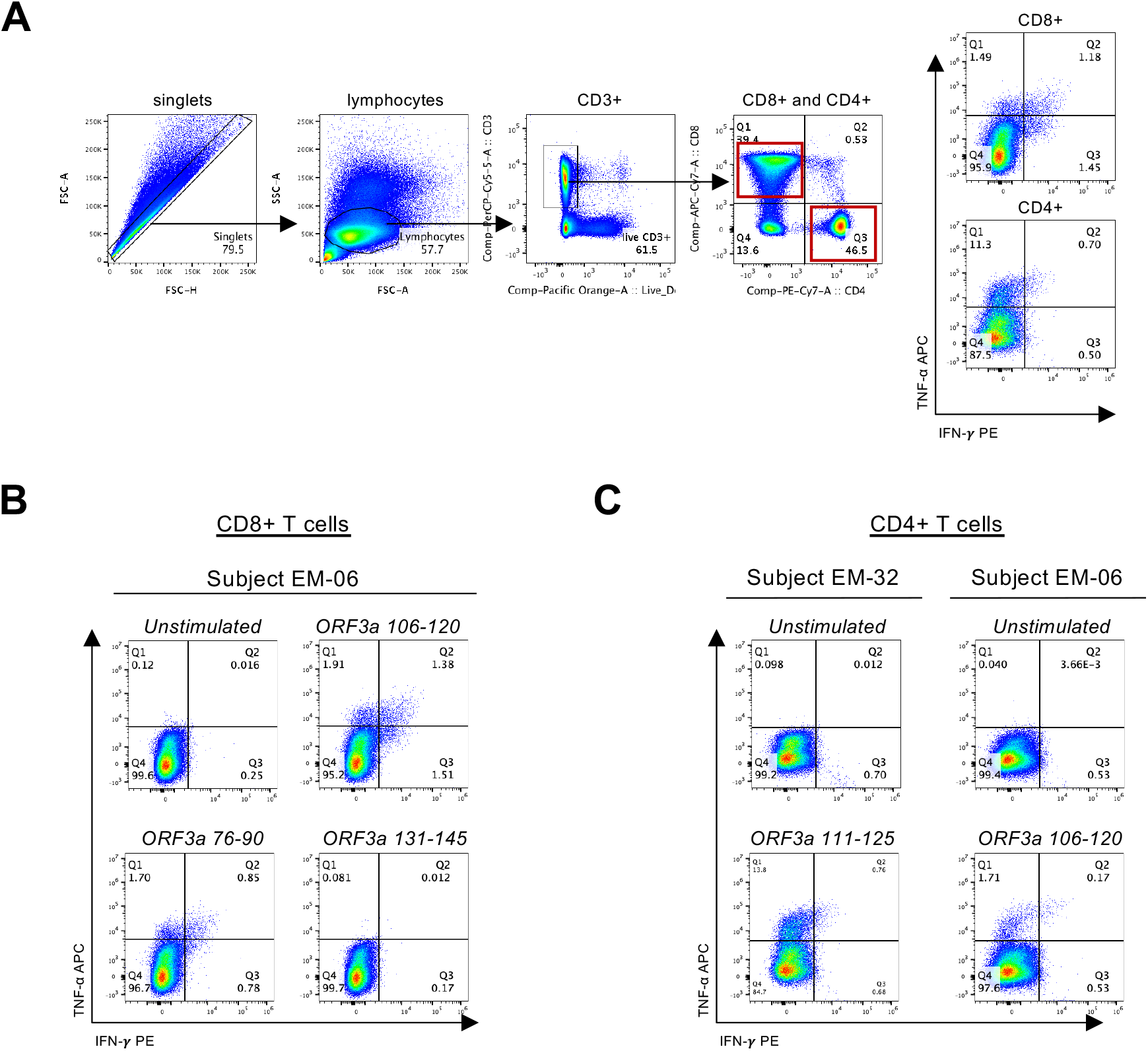
Phenotypic analysis of T cells specific to single SARS-CoV-2 peptides. PBMC were in vitro expanded with peptide pool ORF3a for 10 days and responses to individual peptides were identified stimulating the expanded cells with a peptide matrix strategy using a ELISpot assay. Responses to single peptides were then confirmed by flow cytometry analysis. (**A**) Flow cytometry gating strategy. (**B**) Participant EM-06 with CD8 T cell responses specific to two different peptides within ORF3a. (**C**) Participants EM-32 and EM-06 with CD4 T cell responses specific to two peptides within ORF3a.

## REFERENCES

1. Mbow, M., Lell, B., Jochems, S.P., Cisse, B., Mboup, S., Dewals, B.G., Jaye, A., Dieye, A., and Yazdanbakhsh, M. (2020). COVID-19 in Africa: Dampening the storm? Science 369, 624–626. 10.1126/science.abd3902.

2. Cabore, J.W., Karamagi, H.C., Kipruto, H.K., Mungatu, J.K., Asamani, J.A., Droti, B., Titi-Ofei, R., Seydi, A.B.W., Kidane, S.N., Balde, T., et al. (2022). COVID-19 in the 47 countries of the WHO African region: a modelling analysis of past trends and future patterns. Lancet Global Heal 10, e1099–e1114. 10.1016/s2214-109x(22)00233-9.

3. Njenga, M.K., Dawa, J., Nanyingi, M., Gachohi, J., Ngere, I., Letko, M., Otieno, C.F., Gunn, B.M., and Osoro, E. (2020). Why is There Low Morbidity and Mortality of COVID-19 in Africa? Am J Tropical Medicine Hyg 103, 564–569. 10.4269/ajtmh.20-0474.

4. Ngere, I., Dawa, J., Hunsperger, E., Otieno, N., Masika, M., Amoth, P., Makayotto, L., Nasimiyu, C., Gunn, B.M., Nyawanda, B., et al. (2021). High seroprevalence of SARS-CoV-2 but low infection fatality ratio eight months after introduction in Nairobi, Kenya. Int J Infect Dis 112, 25–34. 10.1016/j.ijid.2021.08.062.

5. Sagar, M., Reifler, K., Rossi, M., Miller, N.S., Sinha, P., White, L., and Mizgerd, J.P. (2020). Recent endemic coronavirus infection is associated with less severe COVID-19. J Clin Invest 131. 10.1172/jci143380.

6. Bartolo, L., Afroz, S., Pan, Y.-G., Xu, R., Williams, L., Lin, C.-F., Tanes, C., Bittinger, K., Friedman, E.S., Gimotty, P.A., et al. (2022). SARS-CoV-2-specific T cells in unexposed adults display broad trafficking potential and cross-react with commensal antigens. Sci Immunol, eabn3127. 10.1126/sciimmunol.abn3127.

7. Habibzadeh, F., Chumakov, K., Sajadi, M.M., Yadollahie, M., Stafford, K., Simi, A., Kottilil, S., Hafizi-Rastani, I., and Gallo, R.C. (2022). Use of oral polio vaccine and the incidence of COVID-19 in the world. Plos One 17, e0265562. 10.1371/journal.pone.0265562.

8. Miyasaka, M. (2020). Is BCG vaccination causally related to reduced COVID-19 mortality? Embo Mol Med 12, e12661. 10.15252/emmm.202012661.

9. Rolot, M., Dougall, A.M., Chetty, A., Javaux, J., Chen, T., Xiao, X., Machiels, B., Selkirk, M.E., Maizels, R.M., Hokke, C., et al. (2018). Helminth-induced IL-4 expands bystander memory CD8+ T cells for early control of viral infection. Nat Commun 9, 4516. 10.1038/s41467-018-06978-5.

10. Uyoga, S., Adetifa, I.M.O., Otiende, M., Yegon, C., Agweyu, A., Warimwe, G.M., and Scott, J.A.G. (2021). Prevalence of SARS-CoV-2 Antibodies From a National Serosurveillance of Kenyan Blood Donors, January-March 2021. Jama 326, 1436. 10.1001/jama.2021.15265.

11. Sekine, T., Perez-Potti, A., Rivera-Ballesteros, O., Strålin, K., Gorin, J.-B., Olsson, A., Llewellyn-Lacey, S., Kamal, H., Bogdanovic, G., Muschiol, S., et al. (2020). Robust T Cell Immunity in Convalescent Individuals with Asymptomatic or Mild COVID-19. Cell 183, 158–168.e14. 10.1016/j.cell.2020.08.017.

12. Le Bert, N., Clapham, H.E., Tan, A.T., Chia, W.N., Tham, C.Y.L., Lim, J.M., Kunasegaran, K., Tan, L.W.L., Dutertre, C.-A., Shankar, N., et al. (2021). Highly functional virus-specific cellular immune response in asymptomatic SARS-CoV-2 infection. J Exp Med 218, e20202617. 10.1084/jem.20202617.

13. Wang, Z., Yang, X., Zhong, J., Zhou, Y., Tang, Z., Zhou, H., He, J., Mei, X., Tang, Y., Lin, B., et al. (2021). Exposure to SARS-CoV-2 generates T-cell memory in the absence of a detectable viral infection. Nat Commun 12, 1724. 10.1038/s41467-021-22036-z.

14. Dan, J.M., Mateus, J., Kato, Y., Hastie, K.M., Yu, E.D., Faliti, C.E., Grifoni, A., Ramirez, S.I., Haupt, S., Frazier, A., et al. (2021). Immunological memory to SARS-CoV-2 assessed for up to 8 months after infection. Science 371, eabf4063. 10.1126/science.abf4063.

15. Group, T.C.-19 C.R.P.S., Herrington, D.M., Sanders, J.W., Wierzba, T.F., Alexander-Miller, M., Espeland, M., Bertoni, A.G., Mathews, A., Seals, A.L., Munawar, I., et al. (2021). Duration of SARS-CoV-2 sero-positivity in a large longitudinal sero-surveillance cohort: the COVID-19 Community Research Partnership. Bmc Infect Dis 21, 889. 10.1186/s12879-021-06517-6.

16. Bertoletti, A., Le Bert, N., and Tan, A.T. (2022). SARS-CoV-2-specific T cells in the changing landscape of the COVID-19 pandemic. Immunity. 10.1016/j.immuni.2022.08.008.

17. Riou, C., Keeton, R., Moyo-Gwete, T., Hermanus, T., Kgagudi, P., Baguma, R., Valley-Omar, Z., Smith, M., Tegally, H., Doolabh, D., et al. (2021). Escape from recognition of SARS-CoV-2 variant spike epitopes but overall preservation of T cell immunity. Sci Transl Med 14, eabj6824.#x2013;eabj6824. 10.1126/scitranslmed.abj6824.

18. Keeton, R., Richardson, S.I., Moyo-Gwete, T., Hermanus, T., Tincho, M.B., Benede, N., Manamela, N.P., Baguma, R., Makhado, Z., Ngomti, A., et al. (2021). Prior infection with SARS-CoV-2 boosts and broadens Ad26.COV2.S immunogenicity in a variant-dependent manner. Cell Host Microbe 29, 1611–1619.e5. 10.1016/j.chom.2021.10.003.

19. Tan, C.W., Chia, W.N., Qin, X., Liu, P., Chen, M.I.-C., Tiu, C., Hu, Z., Chen, V.C.-W., Young, B.E., Sia, W.R., et al. (2020). A SARS-CoV-2 surrogate virus neutralization test based on antibody-mediated blockage of ACE2-spike protein-protein interaction. Nature Biotechnology 395, 470–476. 10.1038/s41587-020-0631-z.

20. Nasimiyu, C., Matoke-Muhia, D., Rono, G.K., Osoro, E., Ouso, D.O., Mwangi, J.M., Mwikwabe, N., Thiong’o, K., Dawa, J., Ngere, I., et al. (2022). Imported SARS-CoV-2 Variants of Concern Drove Spread of Infections across Kenya during the Second Year of the Pandemic. Covid 2, 586–598. 10.3390/covid2050044.

21. Tan, A.T., Lim, J.M.E., Le Bert, N., Kunasegaran, K., Chia, A., Qui, M.D.C., Tan, N., Chia, W.N., Alwis, R. de Ying, D., et al. (2021). Rapid measurement of SARS-CoV-2 spike T cells in whole blood from vaccinated and naturally infected individuals. J Clin Invest 131. 10.1172/jci152379.

22. Ogbe, A., Kronsteiner, B., Skelly, D.T., Pace, M., Brown, A., Adland, E., Adair, K., Akhter, H.D., Ali, M., Ali, S.-E., et al. (2021). T cell assays differentiate clinical and subclinical SARS-CoV-2 infections from cross-reactive antiviral responses. Nat Commun 12, 2055. 10.1038/s41467-021-21856-3.

23. Le Bert, N., Tan, A.T., Kunasegaran, K., Tham, C.Y.L., Hafezi, M., Chia, A., Chng, M.H.Y., Lin, M., Tan, N., Linster, M., et al. (2020). SARS-CoV-2-specific T cell immunity in cases of COVID-19 and SARS, and uninfected controls. Nature 584, 457–462. 10.1038/s41586-020-2550-z.

24. Mateus, J., Grifoni, A., Tarke, A., Sidney, J., Ramirez, S.I., Dan, J.M., Burger, Z.C., Rawlings, S.A., Smith, D.M., Phillips, E., et al. (2020). Selective and cross-reactive SARS-CoV-2 T cell epitopes in unexposed humans. Science, eabd3871. 10.1126/science.abd3871.

25. Braun, J., Loyal, L., Frentsch, M., Wendisch, D., Georg, P., Kurth, F., Hippenstiel, S., Dingeldey, M., Kruse, B., Fauchere, F., et al. (2020). SARS-CoV-2-reactive T cells in healthy donors and patients with COVID-19. Nature 587, 270–274. 10.1038/s41586-020-2598-9.

26. Ford, T., Wenden, C., Mbekeani, A., Dally, L., Cox, J.H., Morin, M., Winstone, N., Hill, A.V.S., Gilmour, J., and Ewer, K.J. (2017). Cryopreservation-related loss of antigen-specific IFNγ producing CD4+ T-cells can skew immunogenicity data in vaccine trials: Lessons from a malaria vaccine trial substudy. Vaccine 35, 1898–1906. 10.1016/j.vaccine.2017.02.038.

27. Grifoni, A., Weiskopf, D., Ramirez, S.I., Mateus, J., Dan, J.M., Moderbacher, C.R., Rawlings, S.A., Sutherland, A., Premkumar, L., Jadi, R.S., et al. (2020). Targets of T Cell Responses to SARS-CoV-2 Coronavirus in Humans with COVID-19 Disease and Unexposed Individuals. Cell 181, 1489–1501.e15. 10.1016/j.cell.2020.05.015.

28. Peng, Y., Mentzer, A.J., Liu, G., Yao, X., Yin, Z., Dong, D., Dejnirattisai, W., Rostron, T., Supasa, P., Liu, C., et al. (2020). Broad and strong memory CD4+ and CD8+ T cells induced by SARS-CoV-2 in UK convalescent individuals following COVID-19. Nat Immunol 21, 1336–1345. 10.1038/s41590-020-0782-6.

29. Zuo, J., Dowell, A.C., Pearce, H., Verma, K., Long, H.M., Begum, J., Aiano, F., Amin-Chowdhury, Z., Hoschler, K., Brooks, T., et al. (2021). Robust SARS-CoV-2-specific T-cell immunity is maintained at 6 months following primary infection. Nat Immunol 22, 620–626. 10.1038/s41590-021-00902-8.

30. Sun, J., Madan, R., Karp, C.L., and Braciale, T.J. (2009). Effector T cells control lung inflammation during acute influenza virus infection by producing IL-10. Nature Medicine 15, 277– 284. 10.1038/nm.1929.

31. Zhao, J., Zhao, J., Mangalam, A.K., Channappanavar, R., Fett, C., Meyerholz, D.K., Agnihothram, S., Baric, R.S., David, C.S., and Perlman, S. (2016). Airway Memory CD4 + T Cells Mediate Protective Immunity against Emerging Respiratory Coronaviruses. Immunity 44, 1379–1391. 10.1016/j.immuni.2016.05.006.

32. Wu, L.-P., Wang, N.-C., Chang, Y.-H., Tian, X.-Y., Na, D.-Y., Zhang, L.-Y., Zheng, L., Lan, T., Wang, L.-F., and Liang, G.-D. (2007). Duration of Antibody Responses after Severe Acute Respiratory Syndrome. Emerg Infect Dis 13, 1562–1564. 10.3201/eid1310.070576.

33. Mok, C.K.P., Zhu, A., Zhao, J., Lau, E.H.Y., Wang, J., Chen, Z., Zhuang, Z., Wang, Y., Alshukairi, A.N., Baharoon, S.A., et al. (2020). T-cell responses to MERS coronavirus infection in people with occupational exposure to dromedary camels in Nigeria: an observational cohort study. The Lancet Infectious Diseases, 1–11. 10.1016/s1473-3099(20)30599-5.

34. Zhao, J., Alshukairi, A.N., Baharoon, S.A., Ahmed, W.A., Bokhari, A.A., Nehdi, A.M., Layqah, L.A., Alghamdi, M.G., Gethamy, M.M.A., Dada, A.M., et al. (2017). Recovery from the Middle East respiratory syndrome is associated with antibody and T-cell responses. Science Immunology 2, eaan5393. 10.1126/sciimmunol.aan5393.

35. Schwarz, M., Torre, D., Lozano-Ojalvo, D., Tan, A.T., Tabaglio, T., Mzoughi, S., Sanchez-Tarjuelo, R., Bert, N.L., Lim, J.M.E., Hatem, S., et al. (2022). Rapid, scalable assessment of SARS-CoV-2 cellular immunity by whole-blood PCR. Nat Biotechnol, 1–10. 10.1038/s41587-022-01347-6.

36. Goletti, D., Petrone, L., Manissero, D., Bertoletti, A., Rao, S., Ndunda, N., Sette, A., and Nikolayevskyy, V. (2021). The potential clinical utility of measuring SARS-CoV-2-specific T-cell responses. Clin Microbiol Infec. 10.1016/j.cmi.2021.07.005.

37. Scurr, M.J., Lippiatt, G., Capitani, L., Bentley, K., Lauder, S.N., Smart, K., Somerville, M.S., Rees, T., Stanton, R.J., Gallimore, A., et al. (2022). Magnitude of venous or capillary blood-derived SARS-CoV-2-specific T cell response determines COVID-19 immunity. Nat Commun 13, 5422. 10.1038/s41467-022-32985-8.

38. Weingarten-Gabbay, S., Klaeger, S., Sarkizova, S., Pearlman, L.R., Chen, D.-Y., Gallagher, K.M.E., Bauer, M.R., Taylor, H.B., Dunn, W.A., Tarr, C., et al. (2021). Profiling SARS-CoV-2 HLA-I peptidome reveals T cell epitopes from out-of-frame ORFs. Cell 184, 3962–3980.e17. 10.1016/j.cell.2021.05.046.

39. Hayn, M., Hirschenberger, M., Koepke, L., Nchioua, R., Straub, J.H., Klute, S., Hunszinger, V., Zech, F., Bozzo, C.P., Aftab, W., et al. (2021). Systematic functional analysis of SARS-CoV-2 proteins uncovers viral innate immune antagonists and remaining vulnerabilities. Cell Reports 35, 109126. 10.1016/j.celrep.2021.109126.

40. Arshad, N., Laurent-Rolle, M., Ahmed, W.S., Hsu, J.C.-C., Mitchell, S.M., Pawlak, J., Sengupta, D., Biswas, K.H., and Cresswell, P. (2022). SARS-CoV-2 accessory proteins ORF7a and ORF3a use distinct mechanisms to downregulate MHC-I surface expression. Biorxiv, 2022.05.17.492198. 10.1101/2022.05.17.492198.

41. Zhang, Y., Chen, Y., Li, Y., Huang, F., Luo, B., Yuan, Y., Xia, B., Ma, X., Yang, T., Yu, F., et al. (2021). The ORF8 protein of SARS-CoV-2 mediates immune evasion through down-regulating MHC-?. P Natl Acad Sci Usa 118, e2024202118. 10.1073/pnas.2024202118.

42. Tan, A.T., Linster, M., Tan, C.W., Le Bert, N., Chia, W.N., Kunasegaran, K., Zhuang, Y., Tham, C.Y.L., Chia, A., Smith, G.JD., et al. (2021). Early induction of functional SARS-CoV-2 specific T cells associates with rapid viral clearance and mild disease in COVID-19 patients. Cell Reports 34, 108728. 10.1016/j.celrep.2021.108728.

43. Ruiter, K. de Jochems, S.P., Tahapary, D.L., Stam, K.A., König, M., Unen, V. van, Laban, S., Höllt, T., Mbow, M., Lelieveldt, B.P.F., et al. (2020). Helminth infections drive heterogeneity in human type 2 and regulatory cells. Sci Transl Med 12. 10.1126/scitranslmed.aaw3703.

44. Mbow, M., Jong, S.E., Meurs, L., Mboup, S., Dieye, T.N., Polman, K., and Yazdanbakhsh, M. (2014). Changes in immunological profile as a function of urbanization and lifestyle. Immunology 143, 569–577. 10.1111/imm.12335.

45. Gleeson, M., Bishop, N.C., Stensel, D.J., Lindley, M.R., Mastana, S.S., and Nimmo, M.A. (2011). The anti-inflammatory effects of exercise: mechanisms and implications for the prevention and treatment of disease. Nat Rev Immunol 11, 607–615. 10.1038/nri3041.

46. Wastyk, H.C., Fragiadakis, G.K., Perelman, D., Dahan, D., Merrill, B.D., Yu, F.B., Topf, M., Gonzalez, C.G., Treuren, W.V., Han, S., et al. (2021). Gut-microbiota-targeted diets modulate human immune status. Cell 184, 4137–4153.e14. 10.1016/j.cell.2021.06.019.

47. Winkler, S., Willheim, M., Baier, K., Schmid, D., Aichelburg, A., Graninger, W., and Kremsner, P.G. (1998). Reciprocal Regulation of Th1- and Th2-Cytokine-Producing T Cells during Clearance of Parasitemia in Plasmodium falciparum Malaria. Infect Immun 66, 6040–6044. 10.1128/iai.66.12.6040-6044.1998.

